# Co-Evaluation of Lactate, Base Excess and Albumin as Predictor of Mortality in Sepsis by Excluding the Factors Which Affect Their Levels

**DOI:** 10.1101/2025.03.23.25324310

**Authors:** Aysenur Gur, Elif Ozturk Ince, Nalan Metin Aksu

## Abstract

**Purpose:** Our aim in this study is to determine the benefits of serum lactate, albumin and base excess values in predicting prognosis and mortality in patients with sepsis or septic shock when evaluated together.

**Methods:** We performed a retrospective observational study. Included 217 patients admitted to Hacettepe University Hospital Adult Emergency Department, that were 18-year-old and more, and had 2 or more SOFA scores. We evaluated admission hour, 24. hour and 48. Hour lactate, albumin and base excess. We searched changes of the lactate, albumin and base excess values and calculate the hospital mortality and 90-days-mortality.

**Results:** Decrease in 0-24-48th hour albumin values increases the mortality of sepsis patients in our study. While 0-48th hour lactate values do not affect the hospital mortality, the increase in the 24th hour lactate value increases the hospital mortality. The increase in 0-48th hour lactate values increases the 90-day mortality. Changes of base excess values had no effect on hospital mortality and 90-day mortality. There was no effect of lactate, albümin and base excess values on mortality in patients with septic shock. When the mortality rates are analyzed according to the lactate clearance of patients with sepsis, hospital mortality increases only as 24-hours lactate clearance decreases. Alactic base excess has no effect on the mortality. While no significant AUC value was found for base excess in ROC analyzes; the AUC values of lactate and albumin are significant, but their sensivity is low since the AUC values found for lactate and albumin are below 0.70. In ROC analyzes for lactate clearance, the AUC value for 24-hour lactate clearance is significant, but the sensivity of the AUC value is low. The areas under the curve (AUC) were not statistically significant in the ROC analyzes for the alactic base excess.

**Conclusions:** Contrary to the literature, lactate, albumin and base excess were found to have low sensitivity in determining prognosis and mortality in our study. When factors that may affect serum lactate, albumin and base excesss (such as chronic liver diseases, chronic kidney diseases, metformin use) are excluded, the values of these biomarkers in determining mortality in sepsis and septic shock decrease.

## 1. INTRODUCTION

Sepsis patients constitute a part of critically ill patients admitted to Emergency Departments(ED). Sepsis is the leading cause of death from infection and a medical emergency [1]. Emergency management of sepsis includes early diagnosis, hemodynamic resuscitation, source control, and appropriate antibiotic administration [2]. It is possible to change the morbidity and mortality results due to sepsis, thanks to appropriate emergency service management.

Sepsis is currently defined as life-threatening organ dysfunction caused by an impaired host response to infection; that is, patients with a SOFA (Sepsis-Related [Sequential] Organ Failure Assessment) score of 2 or higher in patients with suspected infection are defined as septic. Septic shock is the need for a vasopressor to maintain mean arterial blood pressure ≥65 mmHg despite adequate fluid resuscitation and a serum lactate level >2 mmol/L (1).

The SOFA score consists of 6 criterias affecting organ systems (respiratory, cardiovascular, renal, neurological, hepatic and hematological) that are usually measured at admission to the intensive care unit (ICU) and every 24 hours. In recent years, it has been used both in the evaluation of the response of critically ill patients to treatment and in the evaluation of acute morbidity [3].

Although biomarkers add accuracy and objectivity, their usefulness in predicting mortality alone is limited due to their low specificity and sensitivity. Recent studies have shown that the combined use of biomarkers and scoring systems is successful in predicting mortality [4,5]. In some studies, it has been seen that the use of several biomarkers together is beneficial with a scoring system or without a scoring system [6].

Our aim in this study is to determine the benefits of serum lactate, albumin and base excess(BE) values by excluding the factors which affect their levels in predicting prognosis and mortality in patients with sepsis when evaluated together. If we can obtain a meaningful result about these routinely checked biomarkers in critically ill patients, being able to predict the prognosis and mortality of such patients in the ED will benefit us in terms of treatment management.

## 2 MATERIALS and METHODS

Total 217 patients over the age of 18 who applied to the ED, had signs of infection, had a SOFA score of 2 and above, and received written informed consent from themselves or their relatives to participate in the study were included in the study. The study was designed as a prospective observational study. Ethics committee approval with registration number GO 19/510 was obtained from the University Ethics Committee.

Patients’ gender, age, admission complaint, vital signs, SOFA scores on admission, medical history, 0.-24.-48. hour lactate, 0.-24.-48. hour standard BE, 0.-24.-48. hour albumin values, outcomes were recorded.

Septic shock patients were recruited according to the definition in the latest sepsis guideline published by the Surviving Sepsis Campaign in 2016 [1].

SOFA score was calculated according to Table 1.

**Table 1:**
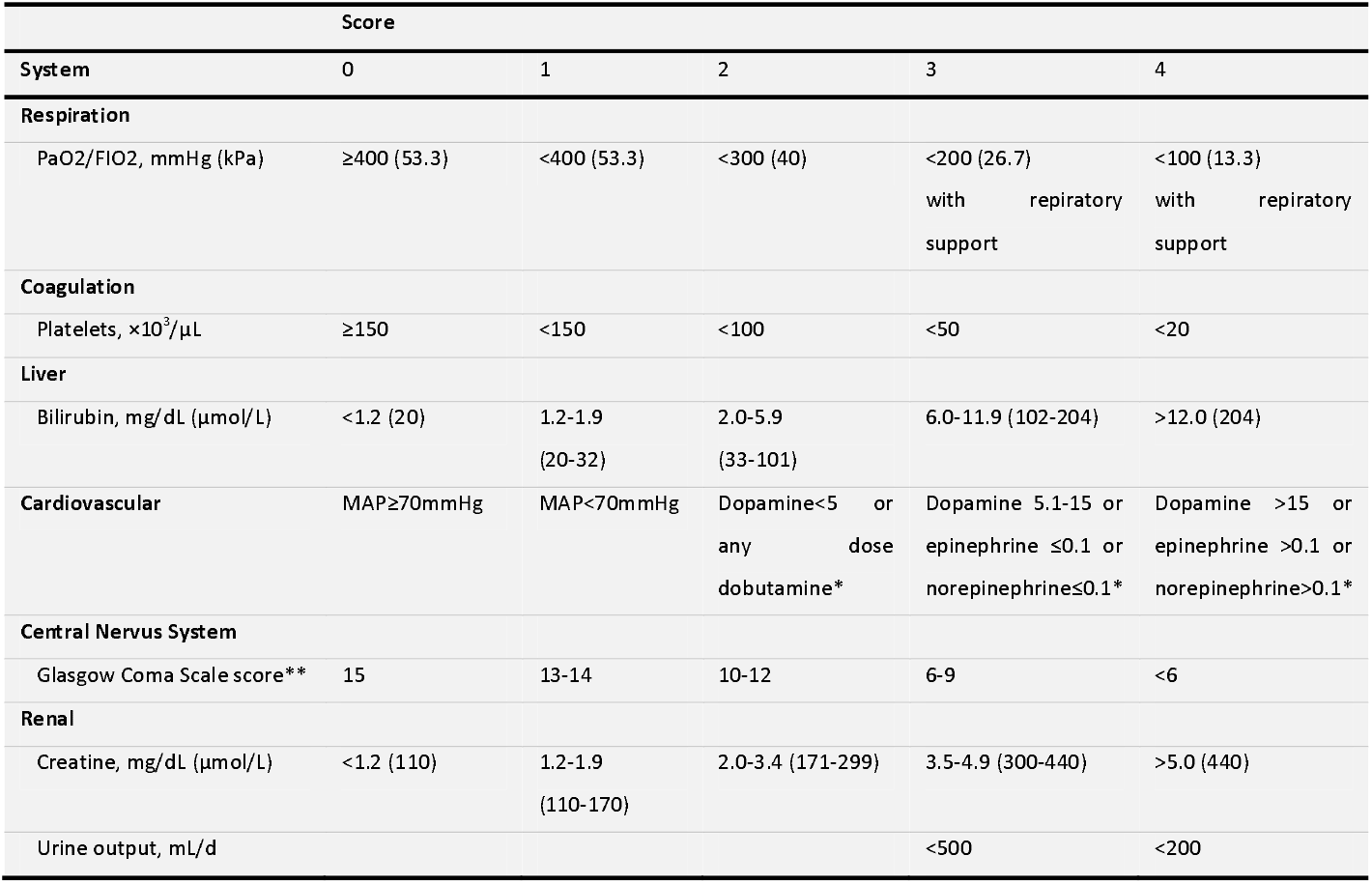
SOFA Score. *Abbreviations: FIO2, fraction of inspired oxygen; MAP, mean arterial pressure; PaO2, partial pressure of oxygen.* *\* Catecholamine doses are given as μg/kg/min for at least 1 hour.* *\*\* GlasgowComa Scale scores range from 3-15; higher score indicates better neurological function*.

The standard BE was calculated according to the formula BE = 0.02786 x pCO2 x 10^(pH - 6.1)^ + 13.77 x pH - 124.58 [7].

The alactic base excess(ABE) was calculated according to the formula ABE (mmol/L) = standard BE (mmol/L) + lactate (mmol/L) [8].

Lactate clearance(LC) was calculated as 24-hour and 48-hour clearance according to the formula LC(%) = (admission lactate-lactate at 24 hours) / admission lactate x100 [9] .

Patients over the age of 18 with signs of infection who presented to the ED and had a SOFA score 2 or higher were included.

Exclusion criterias were as following;

- Being under the age of 18,
- For any reason, lactate, BE or albumin follow-ups are not regular,
- Insufficient information about the outcome could not be reached,
- The admission SOFA score was below 2,
- Type 2 lactatemia reasons (using metformine, chronic liver disease etc.),
- Having chronic kidney disease,
- Antibiotics not started,
- Having acute gastrointestinal bleeding,
- Patients for whom informed consent was not obtained and who did not want to participate in the study,
- Pregnant women,
- Trauma patients,
- Acute myocardial infarction patients,
- Repeated admissions of the same patient

### Stastistical Analysis

Analyzes were performed using SPSS Statistics 23 and the R Shiny application easyROC. Frequency and percentage are given for qualitative variables; mean, standard deviation, median, minimum and maximum values are given for quantitative variables. The relationship between two qualitative variables was examined by Chi-square Analysis. As a result of the chi-square analysis, when the proportion of cells with an expected frequency of less than 5 exceeded 25%, the Final Test results were taken into account, and when it was below 25%, the Pearson Chi-square test results were taken into account. While examining whether there is a difference between the two independent groups in terms of quantitative variables, the assumption of normal distribution was checked first. While checking this assumption, Kolmogorov-Smirnov test of normality and histogram, q-qplot, box-line graphs were used.

“Independent Samples T Test” for the cases where the assumption of normal distribution is provided; in cases where it was not provided, it was applied with the “Mann-Whitney U Test”. Mean and standard deviation values are given for normally distributed variables; median and quartiles are given for non-normally distributed variables. ROC analysis was performed to determine the cut-off value for mortality in terms of 0.-24.-48. hour lactate, BE and albumin variables. In the ROC analysis, firstly, the AUC and the significance of this area were examined. For the variables whose AUC was significant (p<0.05), appropriate cut-off values were determined by looking at the sensitivity and selectivity values. P<0.05 was accepted for statistical significance level. Logistic Regression Analysis was performed in order to evaluate the statistically significant values together.

## 3 RESULTS

The total number of the patients were 217. It was determined that 60.4% of the patients were male (n=131). The mean age of the patients was 67.5±16.5 (the youngest 19, the oldest 98).

Of the patients included in the study, 90.3% (n=196) had sepsis and 9.7% (n=21) had septic shock.

Patients’ 0.-24.-48. hour lactate, LC, BE, ABE and albumin values are analyzed in Table 2.

**Table 2.** Lactate, Lactate Clearance, Base Excess, Alactic Base Excess and Albumin Values.

|  | Mean (Std. Deviation) |
| --- | --- |
| 0. hour lactate* | 2.40 (1.9) |
| 0. hour albumin* | 3 (0.7) |
| 0. hour base excess | -1.47 (6.57) |
| 24. hour lactate* | 1.7 (1.3) |
| 24. hour albumin | 2.65 (0.53) |
| 24. hour base excess | -1.44 (6.04) |
| 48. hour lactate* | 1.5 (1) |
| 48. hour albumin | 2.57 (0.51) |
| 48. hour base excess | -1.3 (6.1) |
| 24-hour lactate clearance* | 26.7 (54.5) |
| 48-hour lactate clearance* | 24 (50) |
| 0. hour alactic base excess | 1.39 (6.25) |
| 24. hour alactic base excess * | 0.2 (7.7) |
| 48. hour alactic base excess | 0.72 (5.56) |

\* The median (IQR) is given for values that do not show normal distribution.

Of the 61.8% (n=134) patients’ infection source were found pulmonary infections. The second source found urinary infections by the ratio 11.5% (n=25).

Of the 32.3% (n=70) died in the hospital. Hospital mortality according to lactate, BE, and albumin values of the patients was analyzed in Table 3. A statistically significant relationship was found between albumin values and hospital mortality. Low albumin values increases hospital mortality. Additionally, the low lactate value in the 24-hour increases the hospital mortality. No statistically significant correlation was found between BE,0.-48. hour lactate values and hospital mortality in sepsis patients. No statistically significant correlation was found between lactate, albumin and BE values and hospital mortality in septic shock patients.

**Table 3.**
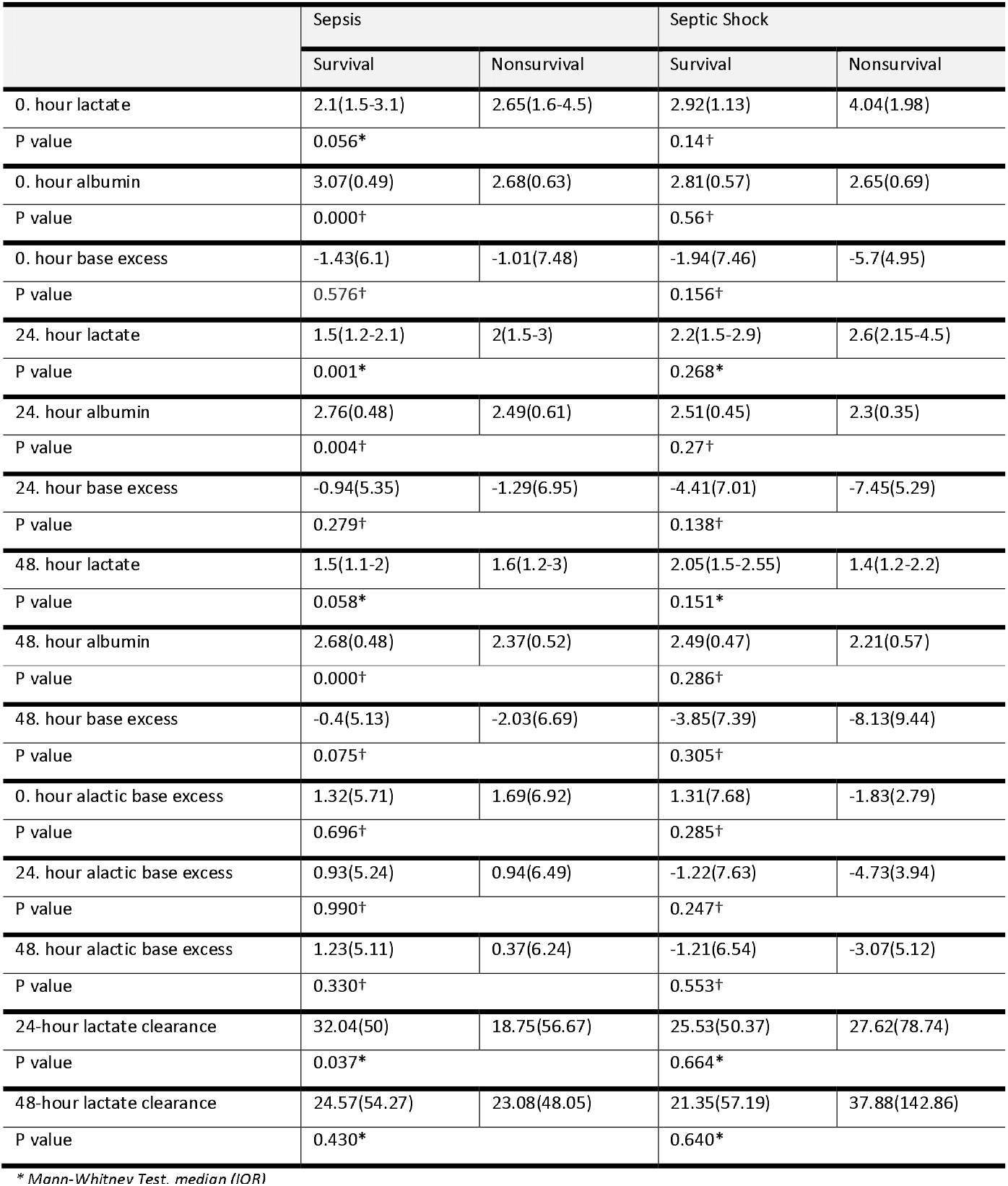
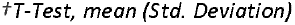
Hospital Mortality according to Lactate, Lactate Clearance, Base Excess, Alactic Base Excess and Albumin Values.

|  | Sepsis |  | Septic Shock |  |
| --- | --- | --- | --- | --- |
|  | Survival | Nonsurvival | Survival | Nonsurvival |
| 0. hour lactate | 2.1(1.5-3.1) | 2.65(1.6-4.5) | 2.92(1.13) | 4.04(1.98) |
| P value | 0.056* |  | 0.14† |  |
| 0. hour albumin | 3.07(0.49) | 2.68(0.63) | 2.81(0.57) | 2.65(0.69) |
| P value | 0.000† |  | 0.56† |  |
| 0. hour base excess | -1.43(6.1) | -1.01(7.48) | -1.94(7.46) | -5.7(4.95) |
| P value | 0.576† |  | 0.156† |  |
| 24. hour lactate | 1.5(1.2-2.1) | 2(1.5-3) | 2.2(1.5-2.9) | 2.6(2.15-4.5) |
| P value | 0.001* |  | 0.268* |  |
| 24. hour albumin | 2.76(0.48) | 2.49(0.61) | 2.51(0.45) | 2.3(0.35) |
| P value | 0.004† |  | 0.27† |  |
| 24. hour base excess | -0.94(5.35) | -1.29(6.95) | -4.41(7.01) | -7.45(5.29) |
| P value | 0.279† |  | 0.138† |  |
| 48. hour lactate | 1.5(1.1-2) | 1.6(1.2-3) | 2.05(1.5-2.55) | 1.4(1.2-2.2) |
| P value | 0.058* |  | 0.151* |  |
| 48. hour albumin | 2.68(0.48) | 2.37(0.52) | 2.49(0.47) | 2.21(0.57) |
| P value | 0.000† |  | 0.286† |  |
| 48. hour base excess | -0.4(5.13) | -2.03(6.69) | -3.85(7.39) | -8.13(9.44) |
| P value | 0.075† |  | 0.305† |  |
| 0. hour alactic base excess | 1.32(5.71) | 1.69(6.92) | 1.31(7.68) | -1.83(2.79) |
| P value | 0.696† |  | 0.285† |  |
| 24. hour alactic base excess | 0.93(5.24) | 0.94(6.49) | -1.22(7.63) | -4.73(3.94) |
| P value | 0.990† |  | 0.247† |  |
| 48. hour alactic base excess | 1.23(5.11) | 0.37(6.24) | -1.21(6.54) | -3.07(5.12) |
| P value | 0.330† |  | 0.553† |  |
| 24-hour lactate clearance | 32.04(50) | 18.75(56.67) | 25.53(50.37) | 27.62(78.74) |
| P value | 0.037* |  | 0.664* |  |
| 48-hour lactate clearance | 24.57(54.27) | 23.08(48.05) | 21.35(57.19) | 37.88(142.86) |
| P value | 0.430* |  | 0.640* |  |

\* Mann-Whitney Test, median (IQR)
<sup>†</sup>T-Test, mean (Std. Deviation)

When mortality rates were analyzed according to ABE and LC in sepsis patients, a statistically significant difference was found only between 24-hour LC and hospital mortality (p=0.037) as shown in Table 4. Hospital mortality increased while 24-hour LC decreased. No significant correlation was found between ABE and LC and the hospital mortality in patients with septic shock.

**Table 4.** Models for Hospital Mortality.

|  | Model-1 |  | Model-2 |  | Model-3 |  |
| --- | --- | --- | --- | --- | --- | --- |
|  | AUC | OR | AUC | OR | AUC | OR |
| 0. hour lactate | 0.517 | 1.143 | - | - | - | 1.116 |
| P value | 0.707 | 0.038 | - | - |  | 0.103 |
| 0. hour albumin | - | - | 0.604 | 0.267 | - | 0.281 |
| P value | - | - | 0.020 | 0.000 |  | 0.103 |
| 0. hour lactate+albumin | - | - | - | - | 0.604 | - |
| P value | - | - | - | - | 0.019 | - |
| 24. hour lactate | 0.570 | 1.635 | - | - | - | 1.580 |
| P value | 0.125 | 0.001 | - | - |  | 0.001 |
| 24. hour albumin | - | - | 0.540 | 0.366 | - | 0.409 |
| P value | - | - | 0.373 | 0.002 |  | 0.008 |
| 24. hour lactate+albumin | - | - | - | - | 0.580 | - |
| P value | - | - | - | - | 0.078 | - |
| 48. hour lactate | 0.588 | 1.610 | - | - | - | 1.516 |
| P value | 0.060 | 0.001 | - | - |  | 0.005 |
| 48. hour albumin | - | - | 0.540 | 0.270 | - | 0.318 |
| P value | - | - | 0.395 | 0.000 |  | 0.002 |
| 48. hour lactate+albumin | - | - | - | - | 0.617 | - |
| P value | - | - | - | - | 0.013 | - |

In order to use lactate, albumin and BE in predicting mortality, models were created by performing ROC analyzes and logistic regression analyzes. Lactate alone in Model-1, albumin alone in Model-2, and lactate and albumin together in Model-3 were evaluated in Table 4. The AUCs were not found statistically significant in the ROC analyzes for BE, ABE and LC as shown in Table 5.

**Table 5.** ROC Analysis of Hospital Mortality for Base Excess, Alactic Base Excess and Lactate Clearance.

|  | Hospital Mortality |  |
| --- | --- | --- |
|  | AUC | P Value |
| 0. hour base excess | 0.500 | 0.995 |
| 24. hour base excess | 0.479 | 0.639 |
| 48. hour base excess | 0.426 | 0.095 |
| 0. hour alactic base excess | 0.510 | 0.837 |
| 24. hour alactic base excess | 0.495 | 0.915 |
| 48-hour alactic base excess | 0.506 | 0.674 |
| 24-hour lactate clearance | 0.412 | 0.047 |
| 48-hour lactate clearance | 0.475 | 0.580 |

When the initial lactate is evaluated alone, it is significant in the model-1 (p=0.038), but the AUC is not significant (p=0.707).

The initial albumin is significant in the model when evaluated alone (p<0.001). Since the odds ratio(OR) is less than 1, the highness in albumin values provides protection. Low albumin values are a risk factor for mortality. The AUC was significant when albumin was used alone (p=0.020); however, since the size of the AUC is below 0.70, its distinctiveness is not high.

When the admission lactate and albumin are used together, the AUC is significant (p=0.019). However, since this model has the same AUC (0.604) with the model in which albumin is alone, it can be said that lactate does not contribute to the model and albumin can be used alone. Since the AUC is below 0.70, the sensitivity values are also low as shown in Table 4.

No significant results were obtained for hospital mortality in the models made according to lactate and albumin values at the twenty-fourth hour.

The lactate+albumin modeling (Model-3) was found to be statistically significant (p=0.013) in the modeling performed according to lactate and albumin values at the 48th hour. However, since the AUC is below 0.70, the sensitivity values are low.

In the ROC analysis for hospital mortality according to the 24-hour and 48-hour LCs of the patients, the AUC for the 24-hour LC was found to be statistically significant (p=0.047); however, the sensitivity value is low because the AUC is below 0.70 as shown in Table 5.

In the ROC analysis performed for hospital mortality according to the patients’ ABE, the AUCs were not statistically significant.

## 4 DISCUSSION

The male patients’ ratio was 60.4 in our study. This rate was consistent with the literature. The mean age of the patients was 67.64±15 years. In the study of Ozaydin et al., the mean age of the patients was 74, in the study of Park et al. was stated as 54 [10,11]. The gender, age and comorbidities of the patients included in studies vary according to the number of patients and the patient profile of the hospitals and regions where studies were conducted.

In our study, the pulmonary sources were found the most common sources of the infection by the ratio 61.8%. In studies of Javed et al. and Park et al., the most common focus of infection was found to be pulmonary respectively (49%), (62%) [12,13] like our study.

We found that 32.3% of the patients died in the hospital. In the study of Freund et al., the hospital mortality was 35% in septic patients with a SOFA score of 2 or higher [15]. It is similar to our mortality rate. In the newly published 2021 sepsis guideline, it is recommended that patients with sepsis or septic shock who need intensive care should be transferred to the ICU within 6 hours [16]; however, in our hospital, patients are admitted to the ICU or service later due to the lack of bed. Despite this, our hospital mortality rates are similar to the literature. At this point, it can be said that our ED has a successful management policy in sepsis management.

It was determined that in our study the low albumin values in 0-24-48 hour increased mortality. 0-48. hour lactate values did not affect the hospital mortality, but it was determined that the high 24th hour lactate value increased the hospital mortality. In the study of Frenkel et al., serum albumin levels on admission were not associated with in-hospital mortality, but one week after admission serum albumin levels were significantly associated with the risk of death in patients with sepsis(p<0.001) [17]. When the lactate values measured at the initial and after 6 hours were examined in the study of Javed et al., lactate values were found to be higher in those who died within 24 hours compared to those who survived (p<0.001) [18]. In the study of Ozaydin et al., initial lactate was examined and 28-day mortality was found to be high in those with high lactate (p=0.007) [14]. Our 24-hour lactate value increases hospital mortality similar to the literature. In the study of Seo et al., it was found that hypoalbuminemia and low BE increased 28-day mortality (p <0.001) [6] . In the study of Lichtenauer et al., it was determined that high initial lactate values (p<0.001) and low initial albumin values (p=0.01) increased mortality [19]. In the study of Yuan et al.,it was found that both too-low (− 41.00 mEq/L to − 2.5 mEq/L) or too-high BE levels (1.9 mEq/L to 55.5 mEq/L) associated with a higher risk of 28-day death for sepsis patients[20]. Low albumin values increase mortality similar to the literature in our study. Contrary to the literature, our BE values were not found to be effective on mortality. This may be related to the fact that the majority of our patients had respiratory problems, and the exclusion of patients with diseases that would affect their basal metabolic status and also albumin and lactate levels, such as chronic kidney disease and chronic liver disease.

In ROC analyzes for lactate, albumin and BE, no significant AUC was found for BE. Since the AUC values determined for lactate and albumin are below 0.70, their discrimination is low. In the study of Lichtenauer et al., the AUC for lactate was 0.804 and the AUC for albumin was 0.755 [19]. This indicates that they have a high distinctiveness in predicting mortality. However, factors affecting lactate and albumin values were not excluded in this study.

In our study, a statistically significant difference was found when mortality rates were analyzed according to 24-hour LC in sepsis patients (p=0.037). Accordingly, as LC decreases at the twenty-fourth hour, hospital mortality increases. In the study of Lee et al., mortality was examined according to 6-hour LC and it was found that a decrease in LC increased mortality (p=0.00) [4]. In the study of Marty et al., mortality was examined according to 6, 12 and 24-hour LCs and it was found that only a decrease in 24-hour LC increased mortality [21]. This is a result similar to our study. In ROC analyzes for LC, the AUC for 24-hour LC is significant, but its discrimination is low because the AUC is below 0.70. Marty et al. found the AUC for 24-hour LC to be 0.79 and the threshold value for LC was determined as -2.1% [22]. In our study, the threshold value could not be determined because the AUC was below 0.70.

In our study, no effect on mortality was determined by ABE values. Therefore, the AUC were not statistically significant in the ROC analyzes for the ABE. The ABE is also an indicator of kidney function and volume status of patients [22]. In our study, patients with chronic kidney disease were excluded and the number of patients receiving vasopressors was small. The fact that our ABE values did not affect mortality may be related to these conditions.

## 5 CONCLUSION

In our study, it was determined that low 0-24-48. hour albumin values, the high 24th hour lactate value and the decrease in the 24-hour LC in sepsis patients increases the hospital mortality on the other hand BE and ABE values didn’t affect hospital mortality. The sensitivity of the values affecting mortality was low; therefore, significant results could not be obtained when evaluated individually or together. The reason why our BE, lactate and albumin values are different from the literature may be that all parameters that would affect these values were excluded from the study in our exclusion criterias and the comorbidities of our patient population.

In limitations; our study was a single-center study. The majority of our patient profile consists of patients with serious comorbidities such as malignancy and frequent hospital admissions. Therefore, regardless of sepsis, the mortality rates of our patients are high due to their current disease.

## Data Availability

All data produced in the present work are contained in the manuscript.

## REFERENCES

1. Singer M, Deutschman CS, Seymour C, et al. The third international consensus definitions for sepsis and septic shock (sepsis-3). Vol. 315, JAMA - Journal of the American Medical Association. American Medical Association; 2016. p. 801–10.

2. Sherwin R, Winters ME, Vilke GM, et al. Does Early and Appropriate Antibiotic Administration Improve Mortality in Emergency Department Patients with Severe Sepsis or Septic Shock? J Emerg Med. 2017;53(4).

3. Lambden S, Laterre PF, Levy MM, et al. The SOFA score - Development, utility and challenges of accurate assessment in clinical trials. Vol. 23, Critical Care. 2019.

4. Lee CC, Chen SY, Tsai CL, et al. Prognostic value of mortality in emergency department sepsis score, procalcitonin, and C-reactive protein in patients with sepsis at the emergency department. Shock. 2008;29(3).

5. Zhao Y, Li C, Jia Y. Evaluation of the mortality in emergency department sepsis score combined with procalcitonin in septic patients. Am J Emerg Med. 2013;31(7).

6. Seo MH, Choa M, You JS, et al. Hypoalbuminemia, low base excess values, and tachypnea predict 28-day mortality in severe sepsis and septic shock patients in the emergency department. Yonsei Med J. 2016;57(6):1361–9.

7. Siggaard-Andersen O. An Acid-Base Chart for Arterial Blood with Normal and Pathophysiological Reference Areas. http://dx.doi.org/103109/00365517109080214. 2009;27(3):p239–45.

8. Gattinoni L, Vasques F, Camporota L, et al. Understanding lactatemia in human sepsis potential impact for early management. Am J Respir Crit Care Med. 2019;200(5).

9. Takahashi N, Nakada T aki, Walley KR, et al. Significance of lactate clearance in septic shock patients with high bilirubin levels. Sci Rep. 2021 Dec 1;11(1).

10. Park M, Pontes Azevedo LC, Maciel AT, et al. Evolutive standard base excess and serum lactate level in severe sepsis and septic shock patients resuscitated with early goal-directed therapy: Still outcome markers? Clinics. 2006;61(1):47–52.

11. Gunes Ozaydin M, Guneysel O, Saridogan F, et al. Are scoring systems sufficient for predicting mortality due to sepsis in the emergency department? 2016; Available from: http://dx.doi.org/10.1016/j.tjem.2016.09.004

12. Kim MH, Choi JH. An update on sepsis biomarkers. Vol. 52, Infection and Chemotherapy. 2020.

13. Javed A, Guirgis FW, Sterling SA, et al. Clinical predictors of early death from sepsis. J Crit Care [Internet]. 2017;42(January 2007):30–4. Available from: https://doi.org/10.1016/j.jcrc.2017.06.024

14. Gunes Ozaydin M, Guneysel O, Saridogan F, et al. Are scoring systems sufficient for predicting mortality due to sepsis in the emergency department? Turkish J Emerg Med. 2017;17(1):25–8.

15. Freund Y, Lemachatti N, Krastinova E, et al. Prognostic accuracy of sepsis-3 criteria for in-hospital mortality among patients with suspected infection presenting to the emergency department. JAMA - J Am Med Assoc. 2017 Jan 17;317(3):301–8.

16. Evans L, Rhodes A, Alhazzani W, et al. Surviving sepsis campaign: international guidelines for management of sepsis and septic shock 2021. Intensive Care Med [Internet]. 2021 Oct 2; Available from: http://www.ncbi.nlm.nih.gov/pubmed/34599691

17. Frenkel A, Novack V, Bichovsky Y, et al. Serum Albumin Levels as a Predictor of Mortality in Patients with Sepsis: A Multicenter Study. Isr Med Assoc J. 2022 Jul;24(7):454-459. PMID: 35819214.

18. Javed A, Guirgis FW, Sterling SA, et al. Clinical predictors of early death from sepsis. J Crit Care. 2017 Dec 1;42:30–4.

19. Lichtenauer M, Wernly B, Ohnewein B, et al. The lactate/albumin ratio: A valuable tool for risk stratification in septic patients admitted to ICU. Int J Mol Sci. 2017;18(9):1–9.

20. Yuan J, Liu X, Liu Y, et al. Association between base excess and 28-day mortality in sepsis patients: A secondary analysis based on the MIMIC-IV database. Heliyon. 2023 May 9;9(5):e15990. doi: 10.1016/j.heliyon.2023.e15990. PMID: 37215834; PMCID: PMC10199177.

21. Marty P, Roquilly A, Vallée F, et al. Lactate clearance for death prediction in severe sepsis or septic shock patients during the first 24 hours in intensive care unit: An observational study. Ann Intensive Care. 2013;3(1):1–7.

22. Respir LGF. Analysis and interpretation: F. Vasques. Am J Respir Crit Care Med [Internet]. 2019 [cited 2022 Jan 17];200(5):582–9. Available from: www.atsjournals.org.

